# An emerging Ferroptosis-Correlated Genes Character for Renal Cell Carcinoma

**DOI:** 10.1101/2023.09.06.23295171

**Authors:** Lin Zhang, Ying Zhou, Jiashun Zhou

## Abstract

Renal Cell Carcinoma (RCC), composed of various subtypes, faces a prognostic prediction challenge. Ferroptosis, an emerging cell death form, has been considered as a vital factor for the development of cancer. However, it remains a further exploration of the predictive value for ferroptosis-correlated genes in RCC. In our work, we paid attention to the gene expression data related to clinical profiles of RCC patients which derived from public databases, TCGA and ICGC. The least absolute shrinkage and selection operator (LASSO) Cox regression performed to establish an optimally predictive model based on the TCGA data. The RCC patients derived from ICGC applied to the validation of the predictive model above. Under expectation, our observation indicated that major ferroptosis-correlated genes (75%) expressed significantly between tumorous and adjacently normal cells in the TCGA cohort. Combined with the differentially expressed genes (DEGs) (P< 0.05) and overall survival (OS), there were twenty-seven genes eventually approached the subsequent analyses. With the optimal 12-genes model screened by LASSO analysis, we aimed to classify RCC patients into the high-risk or low-risk groups based on the median scores. And we created a novel predictive model based on the 12-genes’ risk score. Compared with the two groups, our work discovered that patients in the high-risk group presented more observably shrunken OS than the low (P < 0.05). Through the multivariate cox regression, the predictive model became an independent risk factor in OS (HR =2.053, P<0.001). Moreover, the receiver operating characteristic (ROC) curve proved the significantly predictive value for our predictive model. Functional research unexpectedly showed the immune cells and immune function pathways enriched differently, which emphasized the immune role for the progress of RCC. In a word, an emerging ferroptosis-correlated signature might be popular for medical prediction in RCC. The targeted treatment on ferroptosis may be a potential schedule for RCC.

## 1 Introduction

Renal Cancer, the ninth most common carcinoma around the world[1], accounted for 3% of adult malignancies and had been blamed for 13680 deaths from 65L150 RCC patients in the US in 2013[2]. RCC accounts for 80% of all renal cancers and is famous for late-stage recurrence[3,4]. During the recent two decades, even if the diagnosis and management of RCC were improving, RCC remains one of the most lethally renal malignancies[5]. Smoking, obesity and high blood pressure remain established risk factors for RCC. During 1992 and 2015, one research-based on the SEER database indicated that the incidence of RCC was increasing globally every year[6].

Ferroptosis, an emerging iron-relied pattern of cell death, is activated by the fatal recruitment of lipid peroxidation which significantly increased in the cytoplasm, lipid ROS and mitochondrial membrane density [7,8]. In the recent decade, ferroptosis has become an emerging induction of alternative therapeutics to result in cancer cells’ death, particularly in resistant malignancies[9]. Published studies have illustrated that ferroptosis serves a crucial function in the evolution of RCC. When glutamine and cysteine transformed to glutathione (GSH), ferroptosis accompanied by up-regulated lipid peroxidation[10]. Von Hippel-Lindau (VHL) gene[11], the most frequent genetic event observed in RCC, attracted numerous scholars’ attention for the potential therapeutic in RCC[12–15]. Previous research has shown deprivation of glutamine and cysteine might provide the possibility for VHL/ HIF-related treatment of RCC[16]. Glutamine and cystine were essential for the expression of GSH, which emphasized the deprivation of glutamine and cystine could weaken the expressional GSH accompanied by the alleviated growth in the MYC-dependent RCC[10]. All these shreds of evidence stressed above the relation of ferroptosis-correlated genes and RCC.

In this work, we initially downloaded genetic expression data connected with clinical profiles of RCC patients via public databases TCGA and ICGC. Second, we modelled a ferroptosis-correlated prognostication accompanied with DEGs in the TCGA cohort and verified the predictive value in the ICGC cohort. We established a predictive model based on the relationship between OS and risk. Eventually, our study further attracted to the enrichment of mechanisms to investigate the potential pathway for ferroptosis-correlated genes with RCC.

## 2 Materials and Methods

### 2.1 TCGA-RCC data set and ICGC (RECA-EU) data set

Up to September 24, 2020, we downloaded the gene sequencing data related to clinical information of 1023 RCC patients from the public database the TCGA (https://portal.gdc.cancer.gov/). According to the pathological pattern, we can divide the RCC patients from TCGA into four types such as KIRC, KIRP, KICH and SARC. For the verification of TCGA cohort, the ICGC(https://dcc.icgc.org) cohort provided another 475 tumorous patients. It provided a regulated method in the “limma” R package to average the gene expression value. These prognostic samples primarily derived from US (TCGA), and the verified were from France (ICGC). Because both cohorts came from public databases and obeyed general principle of TCGA and ICGC databases, these observations possessed immunity for approval of local ethics committees. Finally, we retrospected 60 ferroptosis-correlated genes originating from the previous researches [8,9,17–19] in Supplementary Table 1.

### 2.2 Foundation and verification for genetic characteristics of the ferroptosis-prognostic model

Strawberry Perl was used to transforming primary data to the genetic matrix of expression. The “limma” R package played a crucial role to identify the DEGs between tumorous and normal cells with a false discovery rate (FDR) < 0.05 in the TCGA cohort. Combined the clinical information and gene expression, this study explored the ferroptosis-genetic characteristics of OS by univariate Cox analysis. We used Benjamini & Hochberg (BH) correction to adjust P values. The STRING(version 11.0) [20] database originated an interacting genes (IEs) network for the overlapping DEGs and OS. The LASSO-penalized Cox regression analysis, involved in establishing a predictive model, can decrease the statistical risk of over-fitting[21,22]. The R package “glmnet” was used to select optimally genetic variables with LASSO formula. Through the normalized matrix of alternative IEs, we screened the independent variables which contained OS and the alive status of TCGA patients. Patients’ risk scores were calculated with the basement of the adjusted expression value for the corresponding coefficients (CC). The risk-score formula was constructed as follows[19]: score= e^sum^ ^(gene’s^ ^expression^ ^*CC)^. With the median value of the statistical score, the RCC patients classified into high-risk or low-risk groups.

For verification of the prognostic model, PCA was applied into the “prcomp” module via the R package “stats”. Furthermore, the t-SNE analysis was carried out to illustrate the trend of risk-score groups via R package “Rtsne”. The R package “survivalROC” analyzed timeLreliant ROC to test the potential value of the genetic characteristics.

### 2.3 Enrichment analysis

Gene Ontology (GO) and Kyoto Encyclopedia of Genes and Genomes (KEGG) enrichment were constructed by R package “clusterProfiler”. Through the risk-score, several genes were selected to accomplish the immune analysis which contained 16 immune cells and 13 immune pathways with single-sample gene set enrichment (ssGSEA) [23] via the R package “gsva”. Supplementary Table 2 described the infiltrating gene.

### 2.4 Statistical analysis

Student’s t-test was utilized to identify a genetic difference between tumorous and normal cells. The Chi-squared test performed proportions in contrast. Mann-Whitney U test, regulated by the BH analysis, applied to enrich the ssGSEA analysis of immune cells and pathways.

Kaplan-Meier analysis accomplished the OS contrast. To screen independent risk-factors of OS, we adopted univariate and multivariate Cox regression analysis. All statistic accomplished via R software (Version 4.0.0). We take P-value as statistical significance until less than 0.05.

## 3 Results

The establishment of predictive model based on TCGA cohort and verification based on the ICGC cohort just like Fig 1. The present study contained1493 RCC patients derived from 1023 TCGA-RCC cohort, 475 RCC patients derived from ICGC (RECA-EU) cohort. Under the initial screening condition, this work finally involved 881 RCC patients from TCGA and 470 from ICGC. These RCC patients’ clinical information summarized in Table 1.

**Fig 1.**
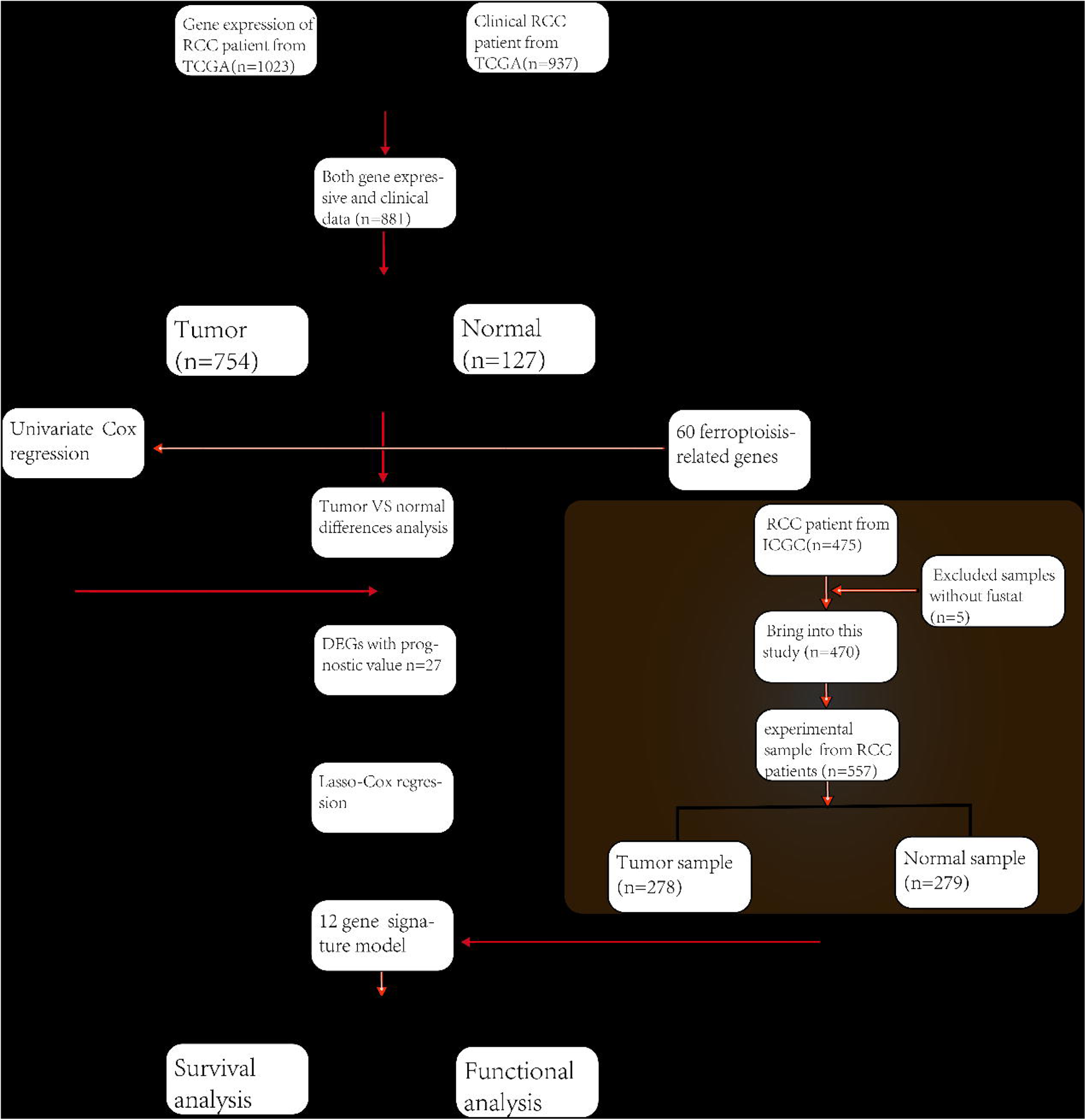
Flow diagram for inclusion and analysis of RCC patients.

**Table 1.** Clinical information of RCC patients from TCGA and ICGC cohort.

|  | TCGA cohort | RECA-EU |
| --- | --- | --- |
| **No. of patients** | 881 | 470 |
| **Age (median, range)** | 60(17-90) | 62(23-86) |
| **Gender (%)** |  |  |
| Female | 288(32.7%) | 194(41.3%) |
| Male | 593(67.3%) | 276(58.7%) |
| **Grade(%)** |  |  |
| Grade 1 | 14(1.6%) | NA |
| Grade 2 | 227(25.8%) | NA |
| Grade 3 | 206(23.4%) | NA |
| Grade 4 | 75(8.5%) | NA |
| Grade 5 | 5(0.6%) | NA |
| unknown | 354(40.2%) | NA |
| **Stage(%)** |  |  |
| I | 456(51.8%) | 226(48.1%) |
| II | 102(11.6%) | 53(11.3%) |
| III | 188(21.3%) | 110(23.4%) |
| IV | 103(11.7%) | 68(14.5%) |
| unknown | 32(3.6%) | 13(2.8%) |
| **Survival status** |  |  |
| OS days (median) | 1071 | 1531 |
| deceased(%) | 226(25.7%) | 137(29.1%) |
| alive(%) | 655(74.3%) | 333(70.9%) |

### 3.1 Recognition of predictive ferroptosis-connected DEGs in the TCGA cohort

Combined ferroptosis-correlated genes with TCGA cohort, we unexpectedly discovered all members of these genes expressed between tumoral and adjacently normal cells in the kidney of RCC. Then the DEGs illustrated that there were 45(45/60,75%) ferroptosis-correlated genes serve a statistically significant function in RCC. According to the OS for the univariate Cox regression analysis, there were 27 genes significantly associated with the clinical data of RCC (Fig. 2a). Compared with the tumoral and normal cells, these 27-genes expressed considerably (Fig. 2b-c). These 27 genes’ correlation network indicated G6PD was the core of the candidate (Fig. 2e).

**Fig 2.**
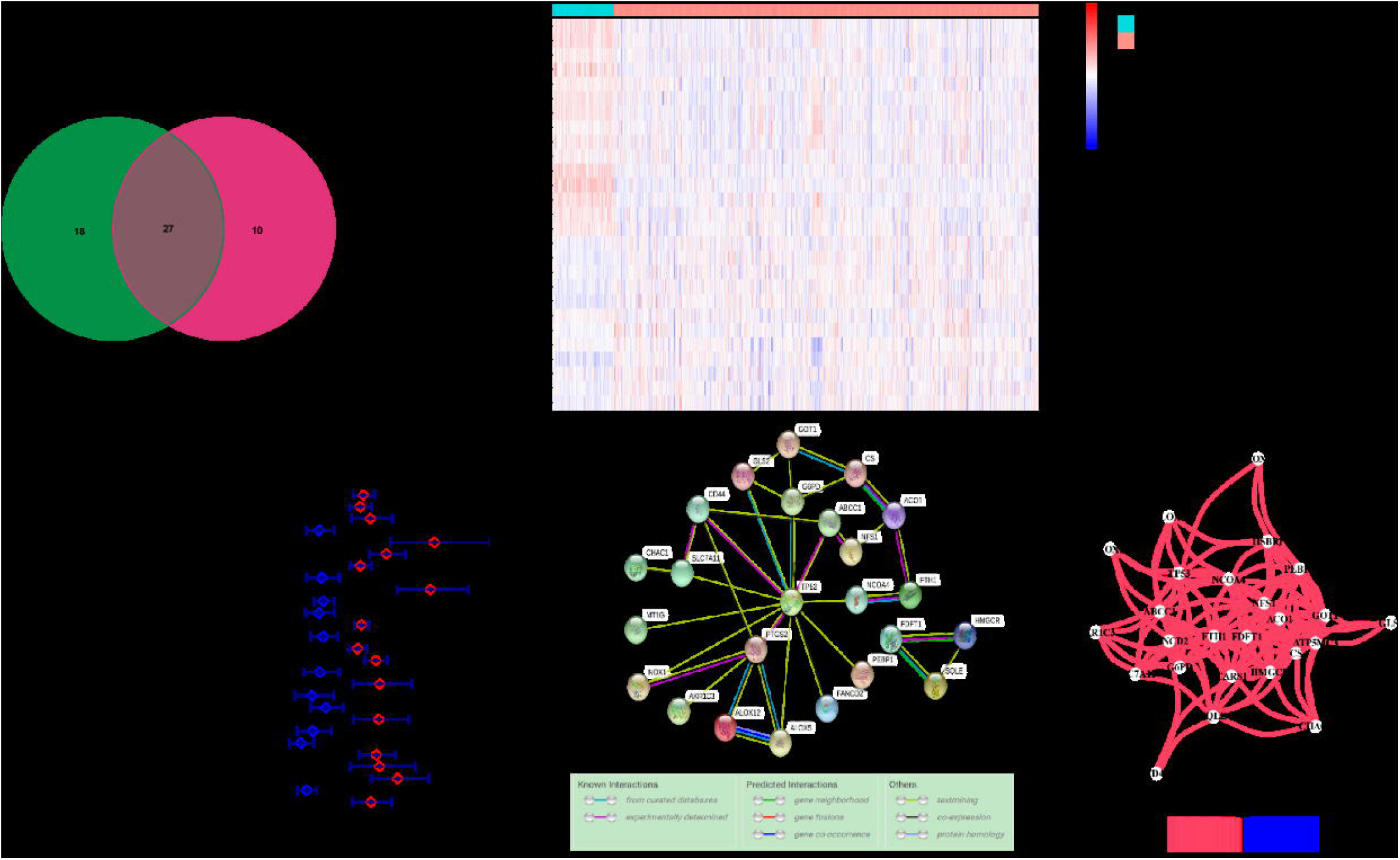
The ferroptosis-correlated genes for RCC patients in the TCGA cohort. a. Venn diagram of the interaction between tumoral and normal tissue for both statistical difference and OS. b. The heat map clustering for 27 genes. c. Forest map illustrates the correlation between the univariate analysis and OS. d. STRING database provided the PPI network to demonstrate the association with the candidate genes. e. The correlation network among candidate genes.

### 3.2 Formation of a predictive model with TCGA cohort

The predictive model, screened from expression profiles of 27 genes mentioned above, constructed a novel risk-score type with LASSO Cox regression analysis. According to the value of λ, our study identified a 12-genes signature composed of seven genes with high expression and five genes with low expression. These 12 genes correlated with survival analysis, and the best cut-off expression value indicated an excellent prognosis (P<0.05). The follows show the formula that we calculated the risk score: e ^(0.043 * expressive value of SLC7A11 + 0.116 * expressive value of G6PD + 0.106 * expressive value of CD44 - 0.004 * expression value of ATP5MC3 + 0.138 * expression value of NOX1 + 0.545 * expression value of FANCD2 + 0.009 * expressive value of PTGS2 - 0.033 * expressive value of ACO1 + 0.136 * expressive value of ABCC1 + 0.159* expressive value of CHAC1 + 0.061 * expressive value of MT1G - 0.301 * expressive value of HMGCR - 0.161 * expressive value^ ^of^ ^GLS2-0.214*^ ^expressive^ ^value^ ^of^ ^PEBP1)^. RCC patients classified into the high-risk group (n=438) or the low-risk group (n=439) with the median cut-off threshold (Fig. 3a). Based on the predictive model, the higher risk group statistically reacted with a higher grade of the tumor. Through t-SNE and PCA analyses, we found patients with different risk-score expressed with different directions (Fig. 3b-c). In the TCGA cohort, high risk-score patients of RCC existed a higher risk of death than the low (Fig. 3d). Similarly, the survival curve showed that patients in the high-risk group possessed more decreased survival time than the low (Fig. 3e, P<0.001). ROC curve, based on the predictively time-dependent model of OS, diagrammatic the area under the curve (AUC) reached 0.755 at one year, 0.739 at two years, and 0.731 at three years (Fig. 3f).

**Fig. 3.**
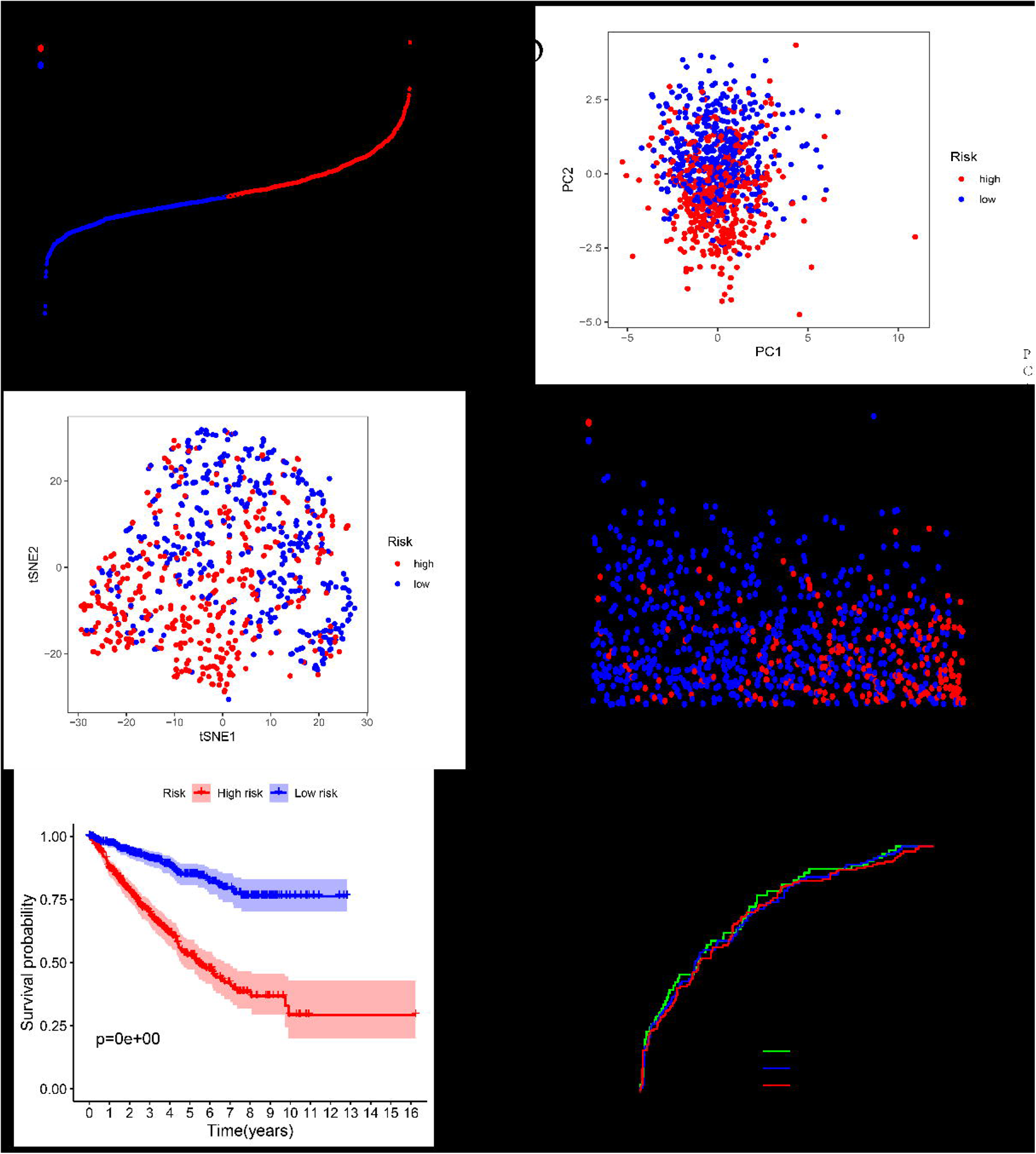
Prognostic model based on the 12-genes in the TCGA cohort. a. The distribution based on a threshold of the risk-scores. b. PCA analysis based on the risk-score groups. c. t-SNE plots based on the risk-score groups. d. The distributions between survival time and increasing risk-score. e. Kaplan-Meier survival curves for the two groups between survival probability and survival time. f. AUC curves tested the predictive value of the prognostic model.

### 3.3 Validation based on the 12-genes in the ICGC cohort

The prognostic model had shown the predictive value for the RCC above. To test the reliability of the model, we also classified RCC patients from ICGC into high-risk or low-risk groups according to the threshold calculated with the similar formula of TCGA cohort. Because of the difference of the two cohorts, this median threshold enacted with the ICGC cohort (Fig. 4a). Consistently, PCA and t-SNE analysis, obtained from the ICGC cohort, showed that patients in different groups distributed in different trends (Fig. 4b-c). The distributions between OS and risk score were not so remarkable like the TCGA cohort (Fig. 4d), but it remained significant. Similarly to the TCGA cohort, the survival curve showed that patients in the high-risk group possessed more decreased survival time than the low (Fig. 4e, P<0.05). Otherwise, the AUC based on the 12-genes was 0.733 at one year,0.645 at two years, and 0.565 at three years (Fig. 4f).

**Fig 4.**
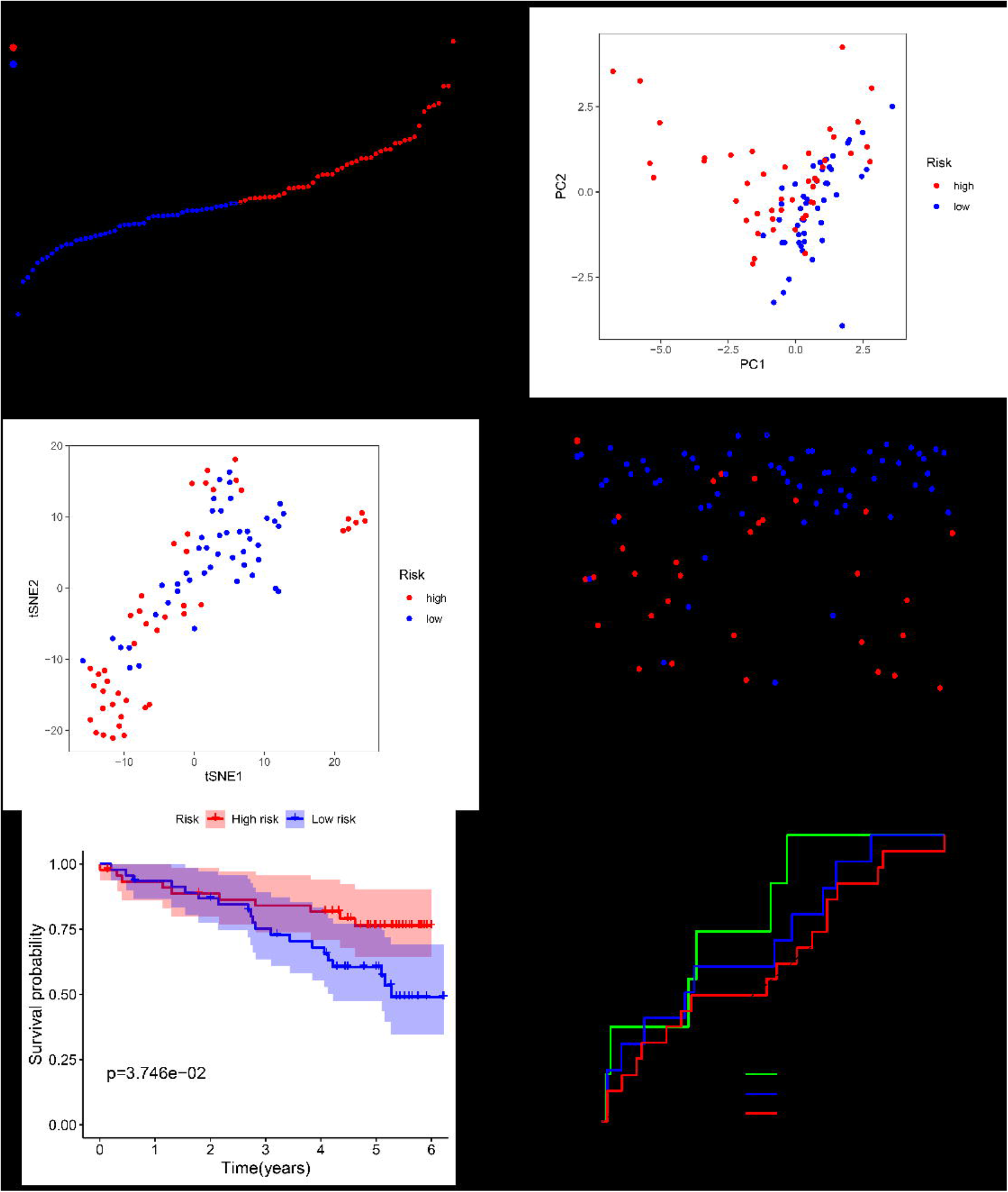
Validation based on the 12-genes in the ICGC cohort. a. The distribution based on a threshold of the risk-scores. b. PCA analysis based on the risk-score groups. c. t-SNE plots based on the risk-score groups. d. The distributions for the two groups between survival time and increasing risk-score. e. Kaplan-Meier survival curves for the two groups between survival probability and survival time. f. AUC of time-dependent ROC curves tested the predictive value of the prognostic model.

### 3.4 Independent predictive value and immune analysis based on the 12-genes in the TCGA cohort

To test whether our predictive model became an independent risk-factor for OS, Cox regression analysis performed the clinical variables. Not only in univariate Cox regression analysis (HR= 2.796, 95% CI =2.252-3.471, P< 0.001 Fig. 5a), but also in multivariate Cox regression analysis (HR =2.053, 95% CI = 1.625-2.592, P<0.001 Fig.5b), our prognostic model had shown that our predictive model closely associated with OS in TCGA cohort.

**Fig 5.**
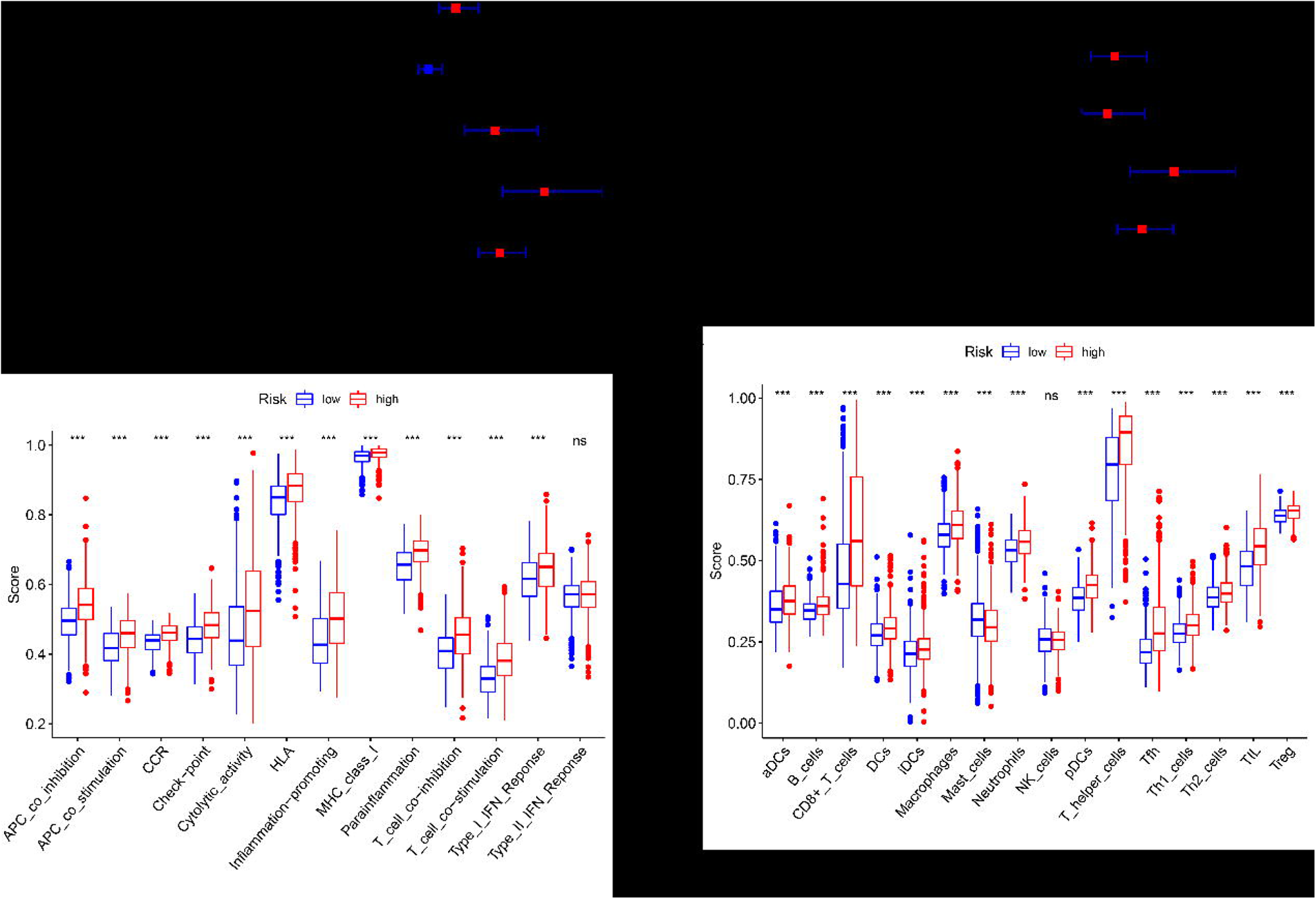
Independent prognostic value and immune analysis of the 12-gene signature. a. Univariate analysis of clinical variables of the TCGA cohort. b. Multivariate analysis of clinical variables of the TCGA cohort. c. Enrichment of various immune pathways. D. Enrichment of various immune cells.

By quantifying the enrichment of various immune cell subpopulations and immune pathways with ssGSEA, we attracted further to investigate the association between the two groups and immune status. Unexpectedly, we found other contents of the immunologic process were significant in the TCGA cohort except for the NK cell and IFN of Type II (adjusted P< 0.05, Fig. 5c-d). Interestingly, the DEGs were also obviously enriched in various and immunity-correlated processes of biology (P. adjust < 0.05, Fig. 5c). We speculated ferroptosis-immune function and cells play an essential role in the development of RCC above.

### 3.5 GO and KEGG analysis both in TCGA and ICGC cohorts

To illustrate the biological functions and pathways which correlated with the predictive model, we dedicated the DEGs between the two groups to perform GO enrichment and KEGG pathway. As expectedly, DEGs involved in two molecularly iron-correlated functions, iron transmembrane transport pathway and iron binding (P. revised< 0.05). In KEGG pathways, there was a common enrichment (protein digestion and absorption pathway) both in TCGA and ICGC cohort (adjusted P< 0.05, Fig. 6c-d).

**Fig 6.**
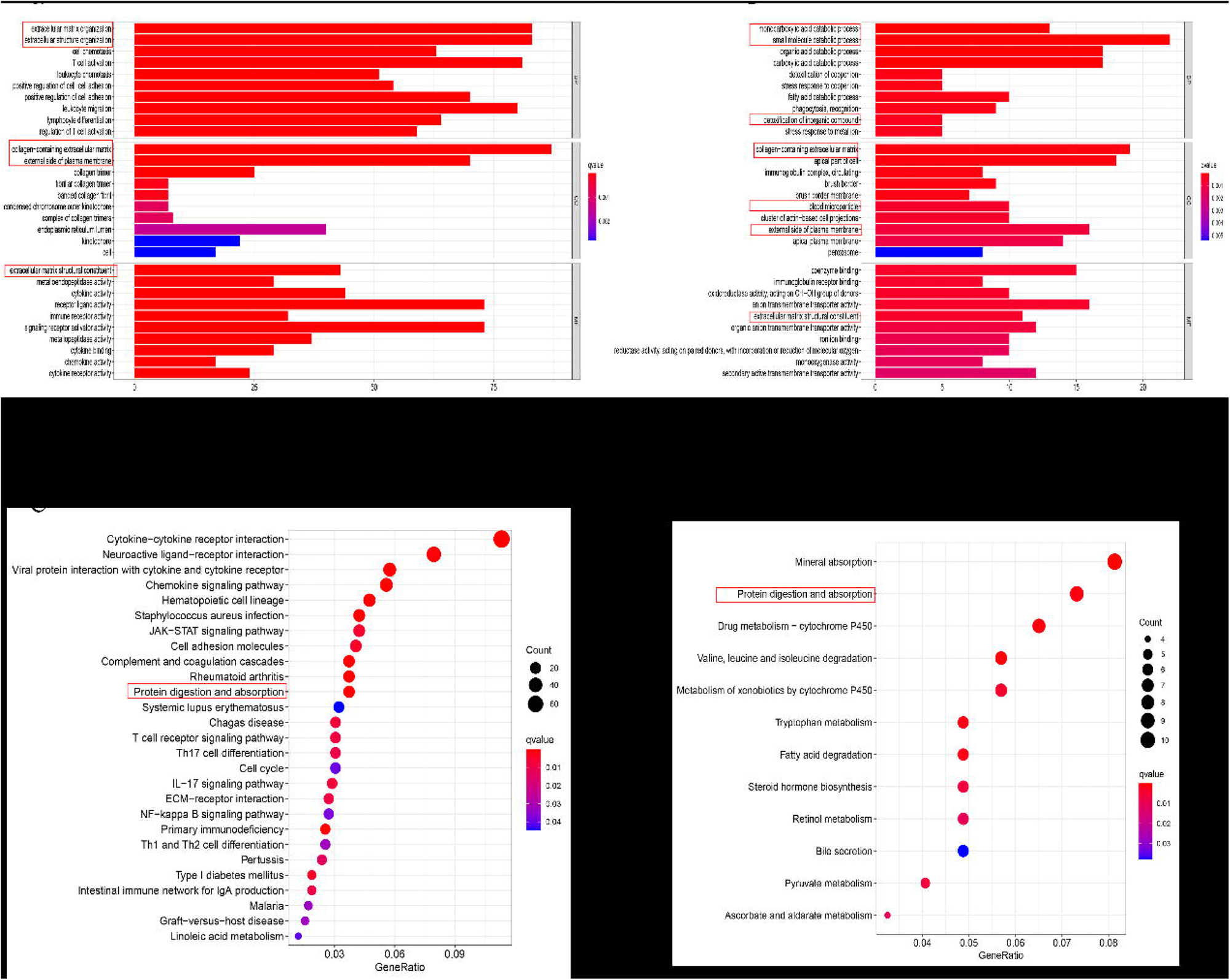
The consequence of enrichment GO (a, b) and KEGG analysis (c, d). The GO enrichment and KEGG pathways in the TCGA cohort (a, c) and ICGC cohort (b, d). The red rectangles show the overlapped enrichments or paths both in the two cohorts.

## 4 Discussion

In this study, we paid attention to investigate the 60 ferroptosis-correlated genes in RCC tumoral cells and their connection with OS. With the LASSO Cox regression analysis to screen a novel predictive model interacting 12 ferroptosis-related genes. To verify whether the predictive model possesses prediction, this work involved an external cohort via the same methods. Functional analysis of GO, KEGG, and immune-related pathways applied to reveal the potential mechanisms of RCC development.

The previous researchs[24] had shown that ferroptosis might serve a crucial function in RCC, but the ferroptosis-correlation with RCC patients’ OS remains unknown. TAZ[25], a hippo pathway effector, triggered a density-regulated ferroptosis through regulating EMP1-NOX4 which indicated a new therapeutic potential for RCC. Compared among 177 types of cancer cells, a recent study shown that RCC is most particularly susceptible to GPX4-regulated ferroptosis[26]. Thus, GPX4’s ferroptosis is an essential regulator of RCC[26]. Sorafenib, a ferroptosis inducer to treat RCC, indirectly supported the existence of ferroptosis in RCC with clinical effectiveness[24].

It amazed us that major ferroptosis-connected genes (75%) significantly expressed compared tumoral with adjacent normal cells, and 27 genes interacted in the Cox regression analysis. Our research showed the potential ferroptosis-target in RCC and the opportunity of creating a predictive model with ferroptosis-correlated genes. This work indicated a prognostic model composed of 12 ferroptosis-correlated genes (ATP5MC3, CD44, NOX1, FANCD2, PTGS2, ACO1, ABCC1, CHAC1, MT1G, HMGCR, GLS2, PEBP1). According to the previous study[9,27], we roughly classified these genes into four categories as follows, lipid metabolism (CD44, PTGS2, HMGCR, PEBP1), iron metabolism (FANCD2, ACO1, MT1G), energy metabolism (ATP5MC3, ABCC1, GLS2) and (anti) oxidant metabolism (NOX1, CHAC1). A meta-analysis reported that CD44 was a prognostic factor in RCC and high CD44 expression connected with high cancer grade, recurrence and poor prognosis[28]. VHL, Vol Hippel-Lindau tumour suppressor gene, accomplished its tumour suppressor action with the restriction of p22phox-based Nox4/Nox1 NADPH oxidase-dependent pathway[29]. GKT137831, a sensitive NOX1/4 isoform inhibitor, observably alleviated ROS and lipid ROS which provided a novel perspective into the molecular-ferroptosis in RMS[30]. FANCD2 and HMGCR [31], reversing the regulation of ferroptosis, were involved in a survival model among five risk-related genes which existed potential prognostic biomarkers and therapeutic to ccRCC[32]. COX-2, regulated by PTGS2, expressed highly in RCC, which might be a trigger that served a crucial function in the development of RCC and warranted further research[33]. ABCC1, as a novel factor of miR-210-3p, negatively regulated miR-210-3p to ease drug-sensitivity of RCC cells. And inhibition of ABCC1 could reverse the effect of miR-210-3p knockdown drug-resistance in RCC cells[34]. PEBP1 can regulate lipoxygenase of lipid death to trigger ferroptosis[35]. Through GCN2-eIF2α-ATF4 pathway, the CHAC1 degradation of glutathione can enhance ferroptosis in breast cancer cells[36]. The published study had proved that MT1G advocated the development of RCC as promoter methylation [37]. It had confirmed that GLS2 play a potential connection of iron-metabolism and ferroptosis to p53tumor suppression[38]. Whether these 12 ferroptosis-genes play an essential role in RCC patients’ prognosis via the development of immune ferroptosis remains to be explained, few researches on these genes except CD44 and ABCC1 had been represented in RCC.

Although it is more and more intense for the ferroptosis-mechanisms of tumour in the recent decade, the potential mechanisms connected tumoral immunity and ferroptosis remains dimness. With the basement of DEGs between high and low risk-score groups, our study applied to GO analysis and amazedly detected that various immune-correlatedly biological pathways enriched. And an expression of different group significantly transformed via immune cells and immune function.

Over the past decade or so, medical treatment for RCC has transformed a nonspecific immune therapy, to targeted medicine against VEGF, and now to an emerging immunotherapy treatment[39] and immune-checkpoint inhibitor have been approved to cure patients with advanced RCC. The tumor immune microenvironment has confirmed to correlate with ccRCC[40]. Tregs and monocytes, both belonged to immune cells, indicated independent risk factors for prognosis in patients with RCC[41].In our study, other types of 15 immune cells and 12 functional immune pathways express significantly in the two groups except for NK cell and the IFN type II. IFN type II independent enhance the up-regulation of B cell TLR7 by NK cells[42], which mean NK cells regulated the IFN type II[43].It was similar to our research, published study illustrated that standard type VHL in human RCC alleviated in NK cells sensitivity[11]. So primary RCC patients expressed no difference in NK cells to IFN type II except the VHL mutated[11].One possible perspective showed that macrophages transported unclear signals, such as lipid factors, recruited antigen-presenting cells (APCs) to the site of ferroptosis [44]. Besides, the high-risk score groups in our study possess a higher possibility of macrophages and Treg cells. As previously described that up-regulated tumoral macrophages [45,46]or Treg cells [47]are correlated with poor prognosis in RCC patients because of immune invasion. Th17 and CD8 cells, Tregs’ subsets, associated positively with improved survival, whereas Th2 cells associated with negative outcomes in ccRCC[47]. The expression of CD4+ and CD25+ regulatory Tregs increased in RCC patients[48], and recombinant IL-2 restrained the number of Tregs in peripheral blood of patients with metastatic RCC via the inhibition of CD4+ and CD25+[49]. In summary, not only our ferroptosis-work but also other studies had confirmed the intense relevance between RCC and immunity. And in our research, there was no different for the NK cells and subsequent IFN type II.

There remained some limitations to our study. Our conclusion required been validated by clinically randomized controlled trials or real-world prospective data because our samples derived from public databases. Second, it is inevitable to ignore some prominent prognostic genes because we merely considered a single signature to build this model of RCC. Importantly, we highlighted the connection for prognostic model and immune activity required experimental verification.

In a word, we reported a novelly predictive model based on 12 ferroptosis-correlated genes which were independently associated with OS in both the TCGA and ICGC cohorts. The potential pathways between ferroptosis-correlated genes and immunity in RCC warranted further investigation.

## Ethics approval

We had waivers of ethics approval because the data came from public database.

## Data Availability

All data produced in the present study are available upon reasonable request to the authors

